# Bridging the rehabilitation data gap in Uganda: a case study exploring the implementation of the WHO Routine Health Information System – Rehabilitation module

**DOI:** 10.1101/2025.02.13.25322214

**Authors:** Md Zabir Hasan, Gerald Okello, Wouter De Groote, Abraham Omaren, Tim Adair, Abdulgafoor M Bachani

**Affiliations:** Johns Hopkins International Injury Research Unit, Department of International Health, Health Systems Program, Johns Hopkins Bloomberg School of Public Health, 615 N Wolfe St, Baltimore, MD 21205, USA; BRAC James P. Grant School of Public Health, BRAC University, 65, BRAC Tower, Floor 10-13, Bir Uttam AK Khandakar Rd, Dhaka 1212, Bangladesh; Makerere University School of Public Health, New Mulago Gate Rd, Kampala, Uganda; Rehabilitation Programme, World Health Organization, 20 Avenue Appia, Geneva, Switzerland; Nossal Institute for Global Health, Melbourne School of Population and Global Health, The University of Melbourne, 207 Bouverie St, Carlton VIC 3053, Australia

**Author notes:** CORRESPONDING AUTHOR: Md Zabir Hasan.

**Keywords:** Rehabilitation, Health information management systems, DHIS2, Implementation Science, Uganda

## Abstract

Uganda’s Health Management Information System (HMIS) has historically lacked robust, standardized data on rehabilitation and assistive technology (AT) services, which has limited effective policy development and service planning. To address this gap, the Ministry of Health sought to integrate the WHO Routine Health Information System (RHIS) – Rehabilitation module into the national digital reporting platform (DHIS2). This mixed-methods explanatory case study adopted the Consolidated Framework for Implementation Research (CFIR) to examine the multi-level processes, determinants, and context influencing integration. The WHO RHIS-Rehabilitation module was integrated through stakeholder engagement, indicator consensus-building, customized tool development, and targeted capacity-building. Six core rehabilitation indicators were prioritized and launched in DHIS2 across 25 referral facilities. The process revealed persistent challenges, including infrastructural constraints, limited workforce capacity, and competing health sector priorities. Nonetheless, substantial improvements were observed in data standardization, stakeholder engagement, and foundational digital reporting capacity for rehabilitation services. Integrating the WHO RHIS–Rehabilitation module into Uganda’s HMIS marks a significant advancement for rehabilitation information systems in resource-limited settings. Key lessons highlight the necessity of early policy alignment, ongoing capacity building, and continuous stakeholder support to sustain and expand rehabilitation data integration. This experience provides a practical pathway for strengthening rehabilitation data systems in comparable contexts.

## 1. INTRODUCTION

The global need for rehabilitation services is increasing because of the rising prevalence of noncommunicable diseases (NCDs), injuries, and aging populations (1). One in three people worldwide lives with a health condition that may require rehabilitation, yet more than 50% of these needs in low– and middle-income countries (LMICs) remain unmet (2). In Uganda, 7.5 million people needed rehabilitation services in 2021, with 26% of the population requiring at least one or more assistive technology (AT) products to support their daily functioning. Only 5% of Ugandans needing an AT product, however, can access it. In comparison, 21% of the population reported unmet needs for AT, reflecting severe deficiencies in availability and affordability, disproportionately affecting women and rural communities (3). These figures indicate the need to expand rehabilitation and AT services, especially in LMICs (4). Investment in rehabilitation and AT enables individuals to achieve and maintain optimal functioning, improve their health outcomes, and enhance their participation in daily life (5).

Establishing a comprehensive and integrated data management system for rehabilitation care is critically important. Such a system is essential to accurately capture all aspects of rehabilitation and AT services, ensuring that data collection is complete, reliable, and accessible for effective planning, monitoring, and improving service delivery. The WHO Rehabilitation 2030: Call for Action emphasizes the importance of strengthening Health Management Information Systems (HMIS) to ensure timely and quality data collection for rehabilitation services (6). This data is critical for defining rehabilitation targets, monitoring outcomes, supporting clinical decision-making, and informing quality management. Routine health facility reporting, integrated into national HMIS, provides essential insights into service availability, distribution, and performance, helping countries achieve Universal Health Coverage (UHC) (7).

Uganda’s national HMIS, based in the District Health Information Software 2 (DHIS2) platform, has historically lacked standardized rehabilitation and AT services indicators (8). This gap has obstructed the systematic documentation of rehabilitation requirements and service delivery, making it challenging for policymakers to prioritize rehabilitation, track service availability, allocate resources effectively, and bridge care gaps. Compounding these challenges, reliance on paper-based data collection further exacerbates data completeness, timeliness, and quality issues. Health facilities face competing demands, limited infrastructure, workforce shortages, and inconsistent reporting practices, particularly in rural and district settings (9).

In response, during the mid-term review of its HMIS, Uganda’s Ministry of Health (MoH) undertook the opportunity to mainstream rehabilitation and AT data within the HMIS, with financial support from USAID, Learning, Acting, and Building for Rehabilitation in Health Systems (ReLAB-HS) Project, and WHO, the MoH collaboratively integrated the WHO’s Routine Health Information System (RHIS) – Rehabilitation module into the national HMIS (10,11) (**Box 1**). Integrating the rehabilitation-specific RHIS module initiative aims to enhance data collection, improve the visibility of rehabilitation services, and inform health policies and strategic resource allocation, enabling Uganda to better respond to increasing rehabilitation needs and ensure equitable, accessible, and quality care (12).

**Box 1:**
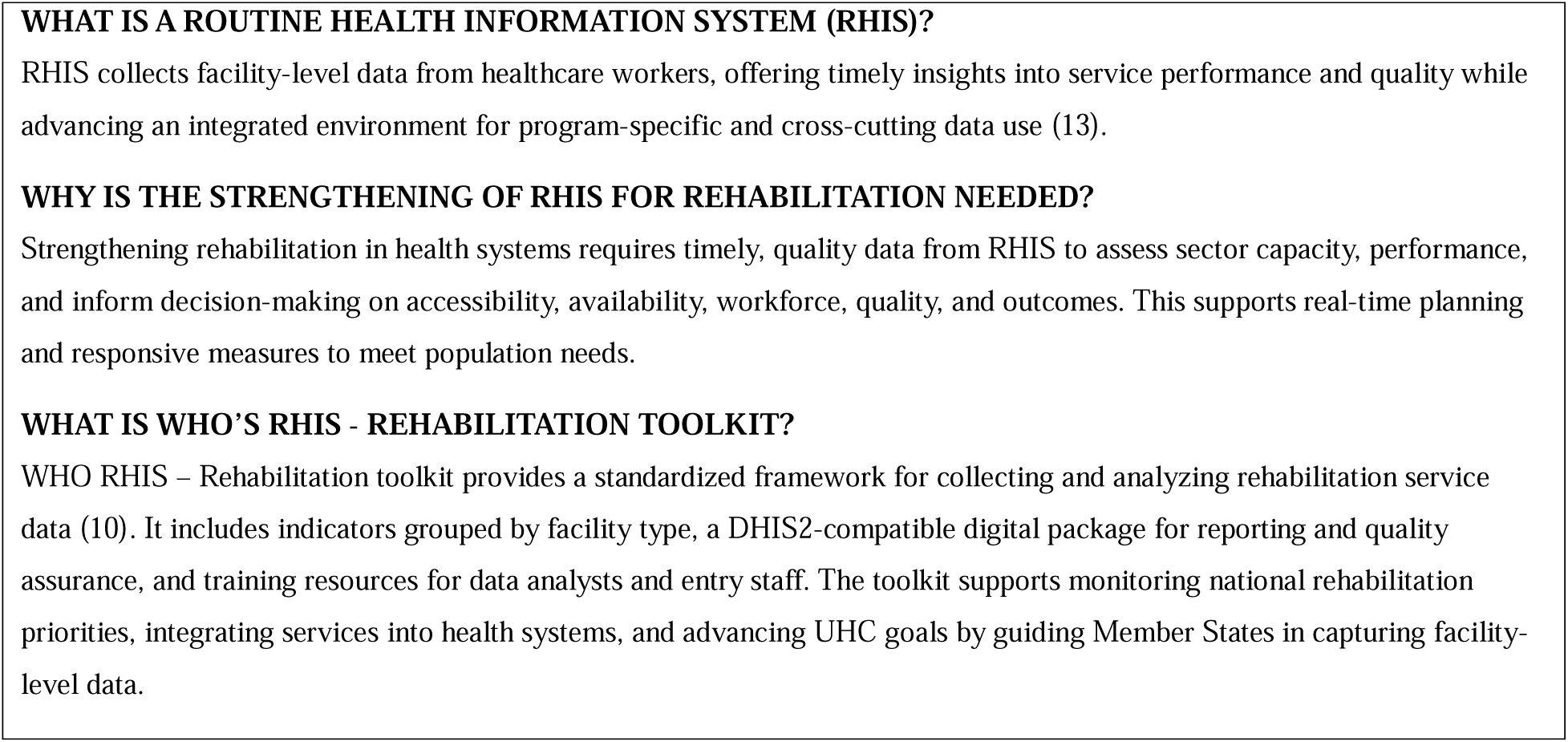
A Snapshot of the Routine Health Information System (RHIS) for Rehabilitation.

This paper examines the integration of the WHO RHIS – Rehabilitation module into Uganda’s national HMIS through an implementation science lens, offering critical insights for strengthening national health information systems in LMICs. Using the Consolidated Framework for Implementation Research (CFIR) (14), we have developed a comprehensive case study that explores the implementation determinants, contextual factors, and implementation processes that facilitated this digital health transformation within Uganda’s DHIS2 platform. Through a detailed examination of the legacy HMIS, the innovation of the RHIS – Rehabilitation module, and the dynamics of both outer and inner settings, as well as implementation processes, we have presented actionable evidence for policymakers, health system planners, and digital health practitioners who seek to strengthen rehabilitation data systems in LMICs. By documenting Uganda’s experience integrating rehabilitation data into its national DHIS2 system, this study contributes novel insights to the growing body of evidence on DHIS2-enabled health systems strengthening, offering practical lessons for countries seeking to mainstream previously neglected primary healthcare (PHC) services into their routine health information systems and ultimately advance progress toward UHC.

## 2. METHODS

### 2.1 Study Design

This study employed a mixed-methods explanatory case study design to examine the integration of the WHO RHIS-Rehabilitation module into Uganda’s national HMIS. Case study methodology is suited for investigating complex, contemporary phenomena within a real-world context, especially when the boundaries between a phenomenon and its context are not clearly evident (15). We selected this approach to understand how and why the integration process unfolded as it did, focusing on the implementation determinants, contextual factors, and strategic processes that influenced this digital health transformation (16). This methodological choice aligns with our theoretical perspective of implementation science, which seeks to understand complex organizational change processes through comprehensive, contextually sensitive approaches that capture multiple perspectives and data sources. The mixed-method explanatory case study design enabled the systematic exploration of the integration process across multiple organizational levels—from national policymakers to facility-level rehabilitation personnel— while focusing on the context of Uganda’s health system (17).

### 2.2 Conceptual Model for Exploring the Integration of the WHO RHIS-Rehabilitation Module

The CFIR served as the primary theoretical framework to guide this case study. It is a meta-theoretical framework, organized across five major domains, all of which determine implementation effectiveness (14,18). This framework is well-suited for examining the complex implementation process because it captures the dynamic interplay between innovation characteristics, outer and inner settings, individual factors, and implementation processes that collectively determine implementation outcomes (19). Additionally, it has demonstrated effectiveness in describing the implementation process of health information systems in similar African contexts, such as Ethiopia, Mozambique, Rwanda, and Zambia (20–22). **Figure 1** illustrates the conceptual framework of this case study, developed based on CFIR domains and their application to the Ugandan context.

**Figure 1:**
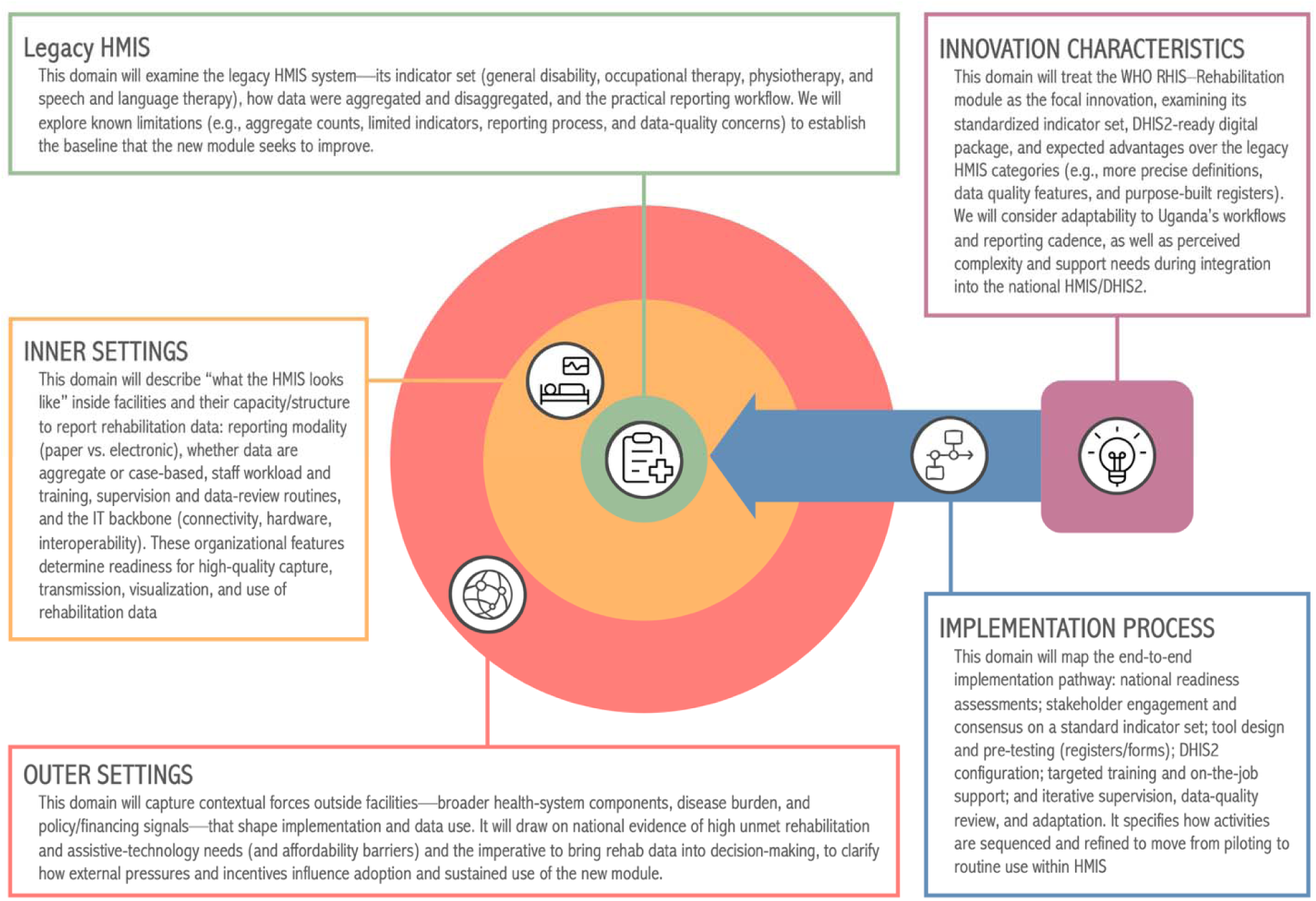
Conceptual framework of the case study based on Consolidated Framework for Implementation Research (CFIR), exploring the integration of the WHO RHIS-Rehabilitation Module into the national health information management system of Uganda.

We adapted the CFIR framework to provide a structured approach for examining implementation determinants across the integration process, maintaining the operational definitions of four core domains while making a strategic modification to enhance the framework’s applicability to our case study context. The Innovation Domain focused on WHO RHIS-Rehabilitation module characteristics, the Implementation Process examined activities and strategies used for integration, the Inner Setting captured facility-level organizational factors, and the Outer Setting encompassed national health policies and external pressures. However, since our case study centered on integrating a digital health innovation into an existing legacy system, we operationalized the Individuals Domain differently than the original framework. Rather than focusing on individual stakeholder roles and characteristics, we conceptualized this domain as representing the Legacy HMIS System of Uganda—the entity receiving and transformed by the WHO RHIS-Rehabilitation module innovation. This adaptation proved valuable for our retrospective mixed-methods approach, which synthesized qualitative interviews, quantitative data, policy documents, and narrative commentary collected over the integration period. By framing the legacy system as the primary “recipient” of the innovation, we could systematically examine how implementation determinants manifested across all CFIR domains and construct a comprehensive understanding of the complex dynamics that influenced the integration process

### 2.3 Data Sources and Analytical Approach

The study drew on both quantitative and qualitative data streams to address the ‘how’ and ‘why’ of the integration process for triangulating findings from diverse perspectives and contexts. We sourced quantitative data from Uganda’s national HMIS using the DHIS2 platform, leveraging routine facility-level reporting. This included aggregated annual and monthly data on rehabilitation and AT indicators from the year 2020-2021. Facility-level registers (both newly introduced and legacy paper-based tools), DHIS2 digital records, and national population projections from the Uganda Bureau of Statistics provided foundational datasets for numerators and denominators required for indicator calculation and analysis. The study further incorporated results from the 2023 Uganda rapid Assistive Technology Assessment (rATA) household survey (3), which provided critical data on population-level needs, access, unmet demand, and barriers for AT. Additional quantitative inputs included results from structured rapid readiness assessments conducted at 12 National and Regional Referral Hospitals and at the national level, as part of the WHO RHIS-Rehabilitation module integration. These assessments collected information on rehabilitation infrastructure, workforce, data capture practices, and digital readiness using standardized questionnaires and WHO-developed checklists.

Qualitative data were derived from multiple complementary sources designed to capture implementation context, stakeholder attitudes, and process complexity. Regular communication and stakeholder consultation with key informants—including MoH officials, facility managers, rehabilitation professionals, HMIS data managers, and healthcare providers—were recorded as meeting minutes, which provided granular insights into readiness, barriers, facilitators, and perceptions of the RHIS-Rehabilitation module integration. Documentation review formed another qualitative evidence base, encompassing policy documents (23), technical guidance notes (8,12), standard operating procedures, records of indicator selection, and tool development workshops (24). These sources highlighted stakeholder engagement chronology, the negotiation and consensus-building processes during indicator adaptation, and real-time challenges and innovations encountered throughout the implementation. The study also utilized narrative comments and reflective logs from trainers and implementing partners, which contextualized quantitative findings and revealed adaptations made in response to emergent constraints (e.g., infrastructure deficits, staff shortages, digital transition challenges, and COVID-19 disruptions).

We synthesized data using an integrative and iterative framework-based approach aligned with the CFIR domains. We conducted descriptive statistical analyses of quantitative facility and national service data and reported on regional disparities, data quality (including completeness, consistency, and timeliness), and service uptake of legacy HMIS indicators. The readiness assessment findings were tabulated and compared across sites to identify patterns of infrastructure gaps, human resource constraints, and IT readiness. The rATA findings helped to contextualize population-level need and system responsiveness. Qualitative materials, including transcripts of group discussions, records, and field notes, underwent thematic content analysis aligned with the CFIR domains (innovation, implementation process, inner and outer setting, and individual characteristics). We developed analytical matrices to examine relationships among determinants, barriers, and facilitators, ensuring that cross-cutting themes and contradictory evidence were adequately captured. We integrated quantitative and qualitative evidence through iterative narrative synthesis, distilling actionable lessons for the Ugandan case and broader health information system strengthening in comparable settings.

## 3. RESULTS

The Results section is organized according to five key domains that shaped the integration of the WHO RHIS-Rehabilitation module into Uganda’s national HMIS. We begin by examining the Legacy HMIS to establish the baseline context and limitations that created the impetus for change, including existing rehabilitation data collection practices and their constraints. The Inner Setting domain focuses on facility-level factors, including organizational characteristics, infrastructure capacity, staff readiness, and local implementation climate. The Outer Setting domain examines macro-level contextual factors—including national health policies, external partnerships, and system-wide pressures—that influenced implementation decisions and sustainability. Next, we described the innovation characteristics of the WHO RHIS-Rehabilitation module, exploring its design features and the adaptations made to its indicator set for Uganda. Finally, the Implementation Process domain details the systematic activities and strategies employed throughout the integration process, from initial planning and stakeholder engagement through training, deployment, and ongoing adaptation.

### 3.1 Legacy HMIS System and Reporting of Existing Rehabilitation Indicators

Uganda’s health information landscape underwent a significant transformation in 2011-12 when the MoH implemented the DHIS2 as its national HMIS, replacing fragmented paper-based reporting mechanisms (25). This transition established a standardized platform for collecting, aggregating, and reporting health data across the country’s health system. Despite this digital advancement, rehabilitation and AT services remained inadequately captured within the national HMIS framework. The reporting system of the facilities captures a limited scope of rehabilitation conditions across four broad categories: general disability (13 conditions), occupational therapy (14 conditions), physiotherapy (17 conditions), and speech and language therapy (7 conditions) (**Table 1**). These data elements represent the rehabilitation-related information currently collected, encompassing a heterogeneous mix of functional conditions (such as difficulty seeing and hearing impairments) and clinical diagnoses (including complex neurological disorders), along with AT device prescriptions. This inconsistent categorization approach complicates systematic data analysis and cross-condition comparisons. Data collection follows standardized demographic disaggregation protocols, with each condition reported by sex and broad age groups (0-28 days, 29 days-4 years, 5-9 years, 10-19 years, 20+ years). However, the aggregated nature of reporting means that data represents total consultation sessions rather than unique individuals, potentially including multiple visits by the same patients throughout the reporting period.

**Table 1:** List of existing rehabilitation-specific indicators in the legacy National Health Information Management System in Uganda.

| No | Data element | Number of conditions included | Example |
| --- | --- | --- | --- |
| 1 | Number of individuals with a specific disability | 13 | Difficulty seeing, difficulty hearing, etc. |
| 2 | Number of individuals with specific physiotherapy conditions | 17 | muscular disorders, joint dysfunction, etc. |
| 3 | Number of individuals with specific physiotherapy conditions who were prescribed assistive devices | 17 | -- |
| 4 | Number of individuals with specific occupational therapy conditions | 14 | Neurodevelopmental Disorders, orthopaedic conditions, etc. |
| 5 | Number of individuals with specific occupational therapy conditions who were prescribed assistive devices | 17 | -- |
| 6 | Number of individuals with specific speech/language therapy conditions | 7 | Speech and language delay/disorder, motor speech disorders, etc. |

Within the existing HMIS architecture, rehabilitation and AT-related data are collected from individual patients during care provision at health facilities and recorded in outpatient registries. These data are subsequently transcribed into paper-based Health Unit Outpatient Monthly Reports at each health facility before being transmitted to the district level, where they are manually entered into the DHIS2 system (**Figure 2**). This multi-step process creates multiple opportunities for data quality deterioration and reporting inconsistencies.

**Figure 2:**
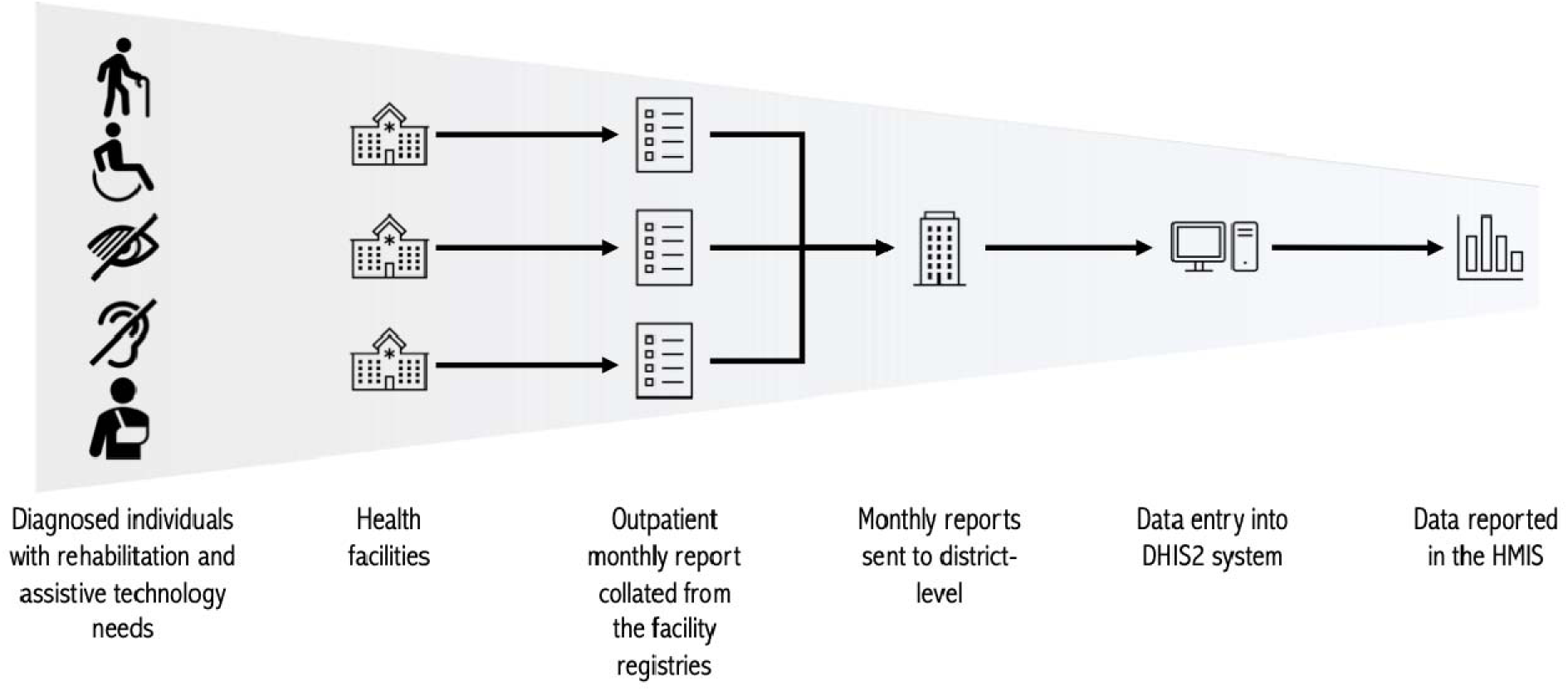
Information Flows in Uganda Health Management Information System.

Analysis of HMIS data from 2020 to 2021 revealed significant systemic challenges undermining the utility of rehabilitation data for health system planning and policy formulation. The data demonstrated consistently low reported utilization of rehabilitation services across all regions, with substantial temporal geographical variations that could not be explained by population demographics or service availability alone. Critical data quality concerns emerged during review of the HMIS data, characterized by inconsistent reporting patterns where health units reported high numbers of specific conditions in one year, followed by significantly fewer cases in subsequent periods (**Figure 3**).

**Figure 3:**
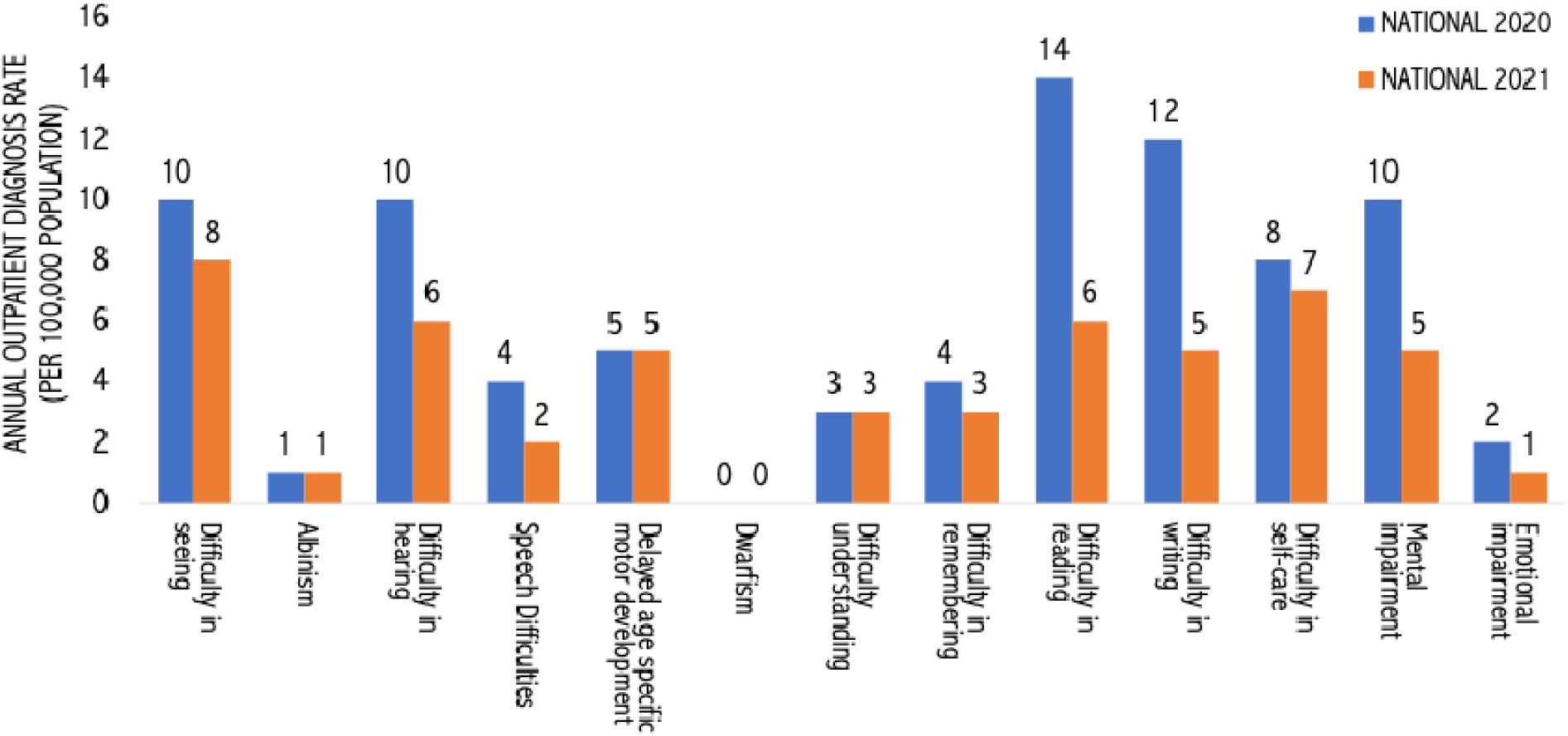
Annual outpatient diagnosis rate (per 100,000 population) reported based on the rehabilitation indicator in the national health information management system in Uganda, 2020–2021.

This erratic reporting pattern suggests fundamental issues with data collection protocols, staff training, or understanding of case definitions, rather than genuine fluctuations in service demand or provision. Moreover, the current indicators fail to offer valuable insights into service utilization, patient outcomes, or specific quality measures, such as outreach activities and waiting times.

### 3.2 Inner Setting of the Health Management Information System

At the facility level, rehabilitation data collection in Uganda operates primarily through fragmented, paper-based systems that lack standardization and integration with the national HMIS. The Rapid Readiness Assessment, conducted across a sample of 12 Regional and National Referral Hospitals in 2023 in preparation for the implementation of the WHO RHIS-Rehabilitation module, revealed that paper-based reporting was widely used at 9 out of 11 sites providing information, while only two facilities—Soroti and Butabika—utilized electronic systems. Individual rehabilitation units collect basic information on patient frequencies and primary conditions, but this data remains largely disconnected from broader facility reporting mechanisms. Staff capacity for data collection and reporting was identified as a critical constraint, with nine sites, excluding Lira and Moroto, reporting inadequate capacity for systematic data collection. The limitations of standardized data collection forms, registers, and indicators for rehabilitation further compound these challenges, as facilities rely on improvised documentation methods that vary significantly across different rehabilitation service units, including physiotherapy, occupational therapy, and orthopedic workshops.

The organizational readiness analysis revealed significant gaps in supervision, training, and data quality assurance mechanisms that directly impact rehabilitation data reporting. Only five facilities—Jinja, Kabale, Moroto, Soroti, and Butabika—had established data quality processes in place, while eight facilities shared data with the MoH, but only three adhered to disease coding standards. The information technology systems support was available at seven sites, with infrastructure components like Local Area Networks (LAN), servers, and internet connectivity provided by the government, though computer availability varied across facilities. Training on data reporting, quality, analysis, and DHIS2 was provided at seven hospitals, but comprehensive HMIS procedures were available at only six facilities. The supervision and monitoring processes showed that seven sites had performance monitoring teams meeting monthly or quarterly.

However, only four used standard records, with barriers including the unavailability of standard records and inadequate staff training on their use. The Systematic Assessment of Rehabilitation Situation (STARS), conducted as part of the MoH report, confirmed these deficiencies, noting fragmented data collection by rehabilitation units lacking collation and consolidation mechanisms (23).

At the national level, the current HMIS architecture presents both opportunities and significant barriers for rehabilitation data integration. The current DHIS2, as its electronic HMIS, is a robust platform that could potentially accommodate rehabilitation data through the newly developed WHO RHIS-Rehabilitation module. However, rehabilitation and AT data are not integrated into either the national HMIS or the complementary Community Health Management Information System (CHMIS), creating a substantial gap in systematic monitoring and evaluation. The STARS indicates that rehabilitation data are not reflected in decision-making and reporting at central policymaking and regional levels, with minimal research in rehabilitation and AT in Uganda. The fragmented approach to data collection by different rehabilitation units, lacking proper collation, consolidation, and appropriate reporting mechanisms, reflects broader systemic challenges in transforming facility-level information into actionable intelligence for policy and resource allocation decisions.

### 3.3 Outer Setting of the Health Management Information System

The broader health system presented some structural challenges that influence the adoption and sustained use of the WHO rehabilitation module within Uganda’s HMIS. Rehabilitation services in Uganda face substantial gaps in accessibility and workforce distribution. Services are primarily concentrated in public National and Regional Referral Hospitals and private health facilities, while district hospitals and lower-tier facilities remain underserved. Of the 1,022 registered rehabilitation professionals—including physiotherapists, occupational therapists, and orthopedic technologists—only 198 were accredited and practicing in the public sector in 2022. The rapid readiness assessment also revealed critical gaps in rehabilitation service delivery infrastructure, with most facilities lacking dedicated rehabilitation wards and operating with severely constrained human resources. Workforce shortages are particularly acute, with speech and language therapists completely absent from all surveyed facilities, audiologists available at only one site (Soroti), and specialized physical medicine and rehabilitation doctors unavailable across the entire network (**Figure 4**). Thus, the current market gap in rehabilitation services is increasingly filled by private providers, non-governmental organizations (NGOs), and faith-based organizations.

**Figure 4:**
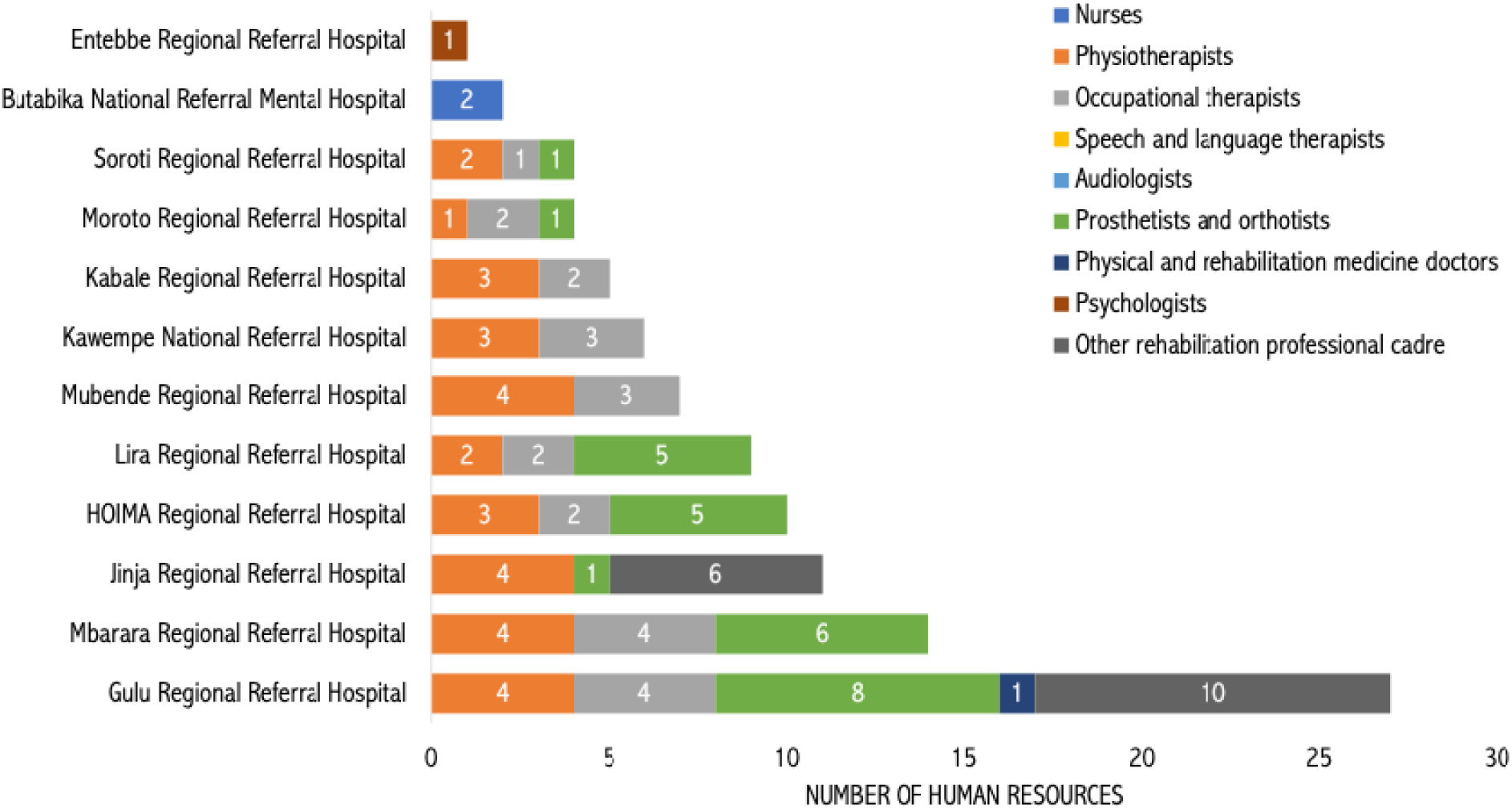
Number of different types of human resources allocated to the rehabilitation department at the National and Regional Referral hospitals, which are included in the rapid readiness assessment.

Health Policy and financing strategies within Uganda’s health system create mixed incentives for rehabilitation data integration, reflecting broader sectoral priorities that have historically marginalized rehabilitation services. The recent STARS revealed that while rehabilitation was positioned within the MoH under the Division of Disability and Rehabilitation, it lacked substantive integration into the Health Sector Development Plan 2021-2024. The plan primarily referenced rehabilitation in the context of physical infrastructure restoration instead of clinical service development. Financing mechanisms presented additional challenges, with rehabilitation services heavily dependent on inconsistent donor support and limited government investment, resulting in high out-of-pocket costs for patients. The absence of a dedicated national rehabilitation strategy or policy framework limited the institutional mandate for systematic data collection and reporting. Also, accountability and transparency mechanisms in the health sector did not encompass all rehabilitation services, making it difficult to track national performance and status. These policy gaps create an environment where rehabilitation data may be perceived as non-essential, potentially undermining staff motivation and organizational commitment to the rigorous data collection protocols required for effective implementation of the WHO RHIS-Rehabilitation module.

Beyond the health sector, it is evident that Uganda faces substantial rehabilitation needs, underscoring the critical importance of establishing robust data systems. However, social and economic barriers significantly constrain access to services. National estimates indicate that approximately one in five Ugandans (7.5 million people) require rehabilitation services, while one in ten experiences functional difficulties requiring rehabilitation and assistive technology. The Rapid Assistive Technology Assessment (rATA) revealed that 26% of Ugandans require at least one assistive product, yet only 5% have access (3). Critically, the sources of AT products demonstrate the limited role of public sector provision, with only 15% of users obtaining devices through public facilities, while 85% access products through NGOs/charities (27%), self-made solutions (25%), friends/family (19%), or private sector providers (14%) (**Figure 5**) (3).

**Figure 5:**
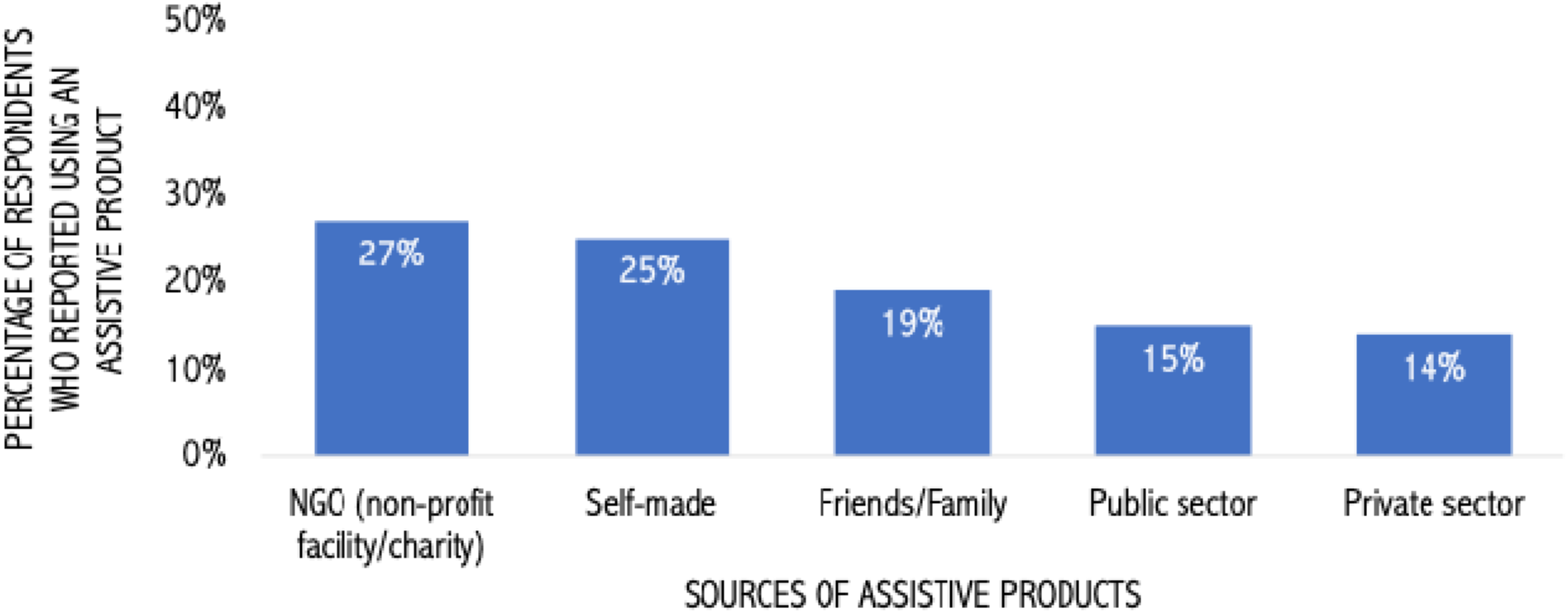
Source of assistive product among those who reported use.

This pattern creates a substantial data blind spot for the HMIS, as the vast majority of rehabilitation and AT interventions outside the government health system remain uncaptured in DHIS2 reporting systems. Moreover, affordability represents the most significant barrier, with 87% of respondents reporting that cost prevents access to AT products, and 21% reporting unmet AT needs. These contextual forces create external pressure for improved rehabilitation data systems as evidence-based advocacy tools, while simultaneously presenting implementation challenges related to low service utilization rates, geographic accessibility constraints, and the complex socioeconomic factors that influence both healthcare-seeking behavior and the capacity of health facilities to systematically document and report rehabilitation service delivery.

### 3.4 The Innovation: WHO RHIS-Rehabilitation module

The WHO RHIS – Rehabilitation module represents a significant advancement over Uganda’s legacy rehabilitation data collection framework. Unlike the existing system’s inconsistent categorization approach that combines heterogeneous data elements across disease and functional categories, the WHO RHIS module provides a standardized, comprehensive framework specifically designed for systematic rehabilitation data capture and analysis. This module comprises 15 indicators: eight core indicators applicable to all health facilities, one for primary care facilities, and six for facilities with dedicated rehabilitation wards (10). These indicators cover critical aspects of rehabilitation services, AT product provision, and patient outcomes (**Table 2**) (26).

**Table 2:** WHO Routine Health Information System – Rehabilitation module indicators.

| Name of the indicator | Numerator | Denominator | Indicator Type | Attributable to |
| --- | --- | --- | --- | --- |
| *All Facilities* |  |  |  |  |
| **Rehabilitation personnel density** | Number of rehabilitation personnel | Total population × 10,000 | Rate (per 10,000 population) | **Assessment of Input** – availability of core resources needed for the provision of essential rehabilitation services, quality of service delivery through the skill set of human resources available in the facilities, distribution of workforce by populations of a catchment area, by level of health care, and by needs specific to a region (equity of workforce deployment), composition of workforce for levels of care and geographical region. |
| **Rehabilitation uptake** | Number of rehabilitation sessions provided | Total population × 10,000 | Rate (per 10,000 population) | **Assessment of Output** – the demand of the population and the capacity of the program to respond (e.g., by health condition group, geography), the extent to which people can access and use rehabilitation services by occupational group (coverage of interventions), and the workload of rehabilitation workers in terms of demand by occupational group. |
| **Rehabilitation service utilization** | Number of cases that receive rehabilitation services | Total population × 10,000 | Rate (per 10,000 population) | **Assessment of Output** – accessibility and availability of services, gender- or age-based accessibility issues, indirect assessment of the quality of services, integration of rehabilitation into a health system (coverage of interventions for specific health condition groups), distribution of services, utilization rates of health condition groups in a selected region or for the same health condition group across regions. |
| **Assistive products uptake** | Number of assistive products provided | N/A | Number (assistive products provided) | **Assessment of Output** – measures rehabilitation accessibility. When disaggregated by the six WHO Assistive Products List categories, this indicator provides information about the capacity to deliver services for the different categories. When disaggregated by geographic region, it describes the capacity of the rehabilitation sector to procure and deliver all assistive product categories nationwide. Also informs on age-based accessibility issues. |
| **Outreach programs uptake** | Number of rehabilitation sessions provided by outreach programs | N/A | Number (rehabilitation sessions provided by outreach programs) | **Assessment of Output** – measures the accessibility of rehabilitation services (such as gender- or age-based accessibility issues) to reduce service barriers. It also reflects rehabilitation demands and the capacity of the health systems to respond. Disaggregation by geographic region can reveal issues by region (distribution of services), with low numbers of sessions provided by outreach programs commonly caused by the lack of availability of required staff and their transport. |
| **Rehabilitation referral** | Number of referrals × 100 | Total number of new cases accessing the facility | Percentage | **Assessment of Efficiency** – assessment of the continuum of care and, therefore, the quality and effectiveness of care. This can be used as a proxy indication for effective referral practice at all levels of care, when measured by geographic region can help establish or improve referral systems for rehabilitation or for awareness raising and training purposes, at the tertiary level can measure effective referral for people with complex rehabilitation needs, at primary health care level can demonstrate continued rehabilitation for chronic conditions and follow up closer to the community. Assessment of referrals by service type. |
| **Rehabilitation waiting time** | Total of waiting days for the first session | Total number of new cases | Average (waiting days) | **Assessment of Efficiency** – informs the estimation of unmet needs and timely service delivery. Analysis by occupational group for different levels of healthcare helps identify the need for investments regarding the availability of the rehabilitation workforce at various levels. Analysis by region can aid in advocacy efforts to secure additional personnel, and comparison by facility can benefit facility managers. |
| **Waiting time for assistive product provision** | Total of waiting days for assistive product provision | Total number of assistive products provided | Average (waiting days) | **Assessment of Efficiency** – assessment of the continuum of care; timeliness of service delivery. It can measure whether there are delays, which can indicate poor capacity for service delivery or problems with procurement processes. Waiting times may differ by WHO Assistive Products List categories. These can be used to assess interventions to reduce waiting times and compare facilities of the same level of healthcare. It also assesses accessibility issues in terms of unmet needs for different APL categories and for different geographical regions. |
| **Primary care facilities** |  |  |  |  |
| **Essential package availability** | Number of facilities offering an Essential Package for rehabilitation × 100 | Total number of facilities | Percentage | **Assessment of Output** – measures the percentage of PHC facilities providing one or more essential recommended basic packages and includes information on the status of the implementation of this recommendation in countries. This contributes to the assessment of Universal Health Coverage. Can be disaggregated by type of essential package, which informs the availability of service types. Analysis by geographical region can inform distribution equality. |
| **Dedicated rehabilitation ward** |  |  |  |  |
| **Rehabilitation bed density** | Number of rehabilitation beds | Total population × 10,000 | Rate (per 10,000 population) | **Assessment of Input** – availability of core resources needed for the provision of essential rehabilitation services, monitoring, planning, and coordination, i.e., distribution of rehabilitation beds across health facilities and across programs to meet the needs of target populations. Indirectly assesses the quality of the service delivery system. Informs readiness for changing rehabilitation needs, especially in disaster-prone countries. |
| **Individualized care plan** | Number of new inpatients receiving an individualized care plan × 100 | Total number of new inpatients | Percentage | **Assessment of Output** – This is a measure of quality for dedicated inpatient rehabilitation services because an individualized care plan contributes to better rehabilitation outcomes for clients. Analyzing this indicator over time provides insight into the progress made toward implementing individualized care plans for all clients, which can be compared across hospitals and regions. |
| **Length of stay** | Number of days of inpatient stay (for discharged clients) | Number of discharges | Average (admission days) | **Assessment of Output** – This is a measure of quality for dedicated inpatient rehabilitation services, as the length of a rehabilitation stay informs assessment of effectiveness and efficiency. Benchmarking for a length of stay is challenging, but the aim should be to achieve a shorter length of stay while respecting the client's progress in improving their functioning. An assessment of the length of stay for a specific health condition over a period informs about the progress made toward improvement of the effectiveness of rehabilitation services for each health condition, which may result from interventions. This indicator also informs rehabilitation service use for selected health conditions. |
| **Functioning change** | Difference between the clients' average functioning assessment score at admission and at discharge | N/A | Difference | **Assessment of Outcome** – measures the change in the ability to independently live and participate in society, which is the overall aim of rehabilitation, it can assess overall clinical outcomes of rehabilitation and inform policymakers about the results of rehabilitation, assessment by facility can be conducted for quality assurance purposes and also as evidence for the planning of quality improvement measures, provides insights into achievable rehabilitation goals for that particular health condition |
| **Coverage for people with acute and complex needs** | Number of first-time admissions for selected health conditions × 100 | Estimated number of new cases for selected health conditions in the country | Percentage | **Assessment of Outcome** – Assesses good clinical practice (prescribing inpatient multi-disciplinary rehabilitation in a dedicated rehabilitation ward) for the effective rehabilitation of a group of health conditions with acute and complex rehabilitation needs. Provides evidence of access issues and/or perceptions of service quality (acceptability). The choice of health condition to be measured should be based on national priorities and the availability of the denominator (i.e., the estimated number of new cases in the country with a defined health condition). Hence, this indicator can only be captured at (sub)national level. The denominator could be a national estimate from ICD coding, or incidence rates for the region can be used to calculate country-specific denominators. This indicator also indirectly assesses the quality of rehabilitation services and the effectiveness of rehabilitation referral practices. |
| **Accessibility for people with acute and complex needs availability** | Number of first-time admissions for a selected health condition | N/A | Number | **Assessment of Outcome** – Dedicated rehabilitation ward: Can be used where a denominator is unavailable for 'Coverage for people with acute and complex needs.' This indicator is mapped to show locations of the specialized facilities with dedicated rehabilitation wards, comparing their overall utilization data and rehabilitation bed density, which can also be assessed by specific health conditions and geographically (assessing equitable access across health conditions and regional capacity issues). |

This DHIS2-compatible configuration package facilitates seamless integration with existing national health information platforms, while purpose-built data collection registers ensure consistent documentation protocols across facilities. Built-in data quality assurance features include validation rules, automated outlier detection, and consistency checks. The standardized analysis framework enables cross-facility comparisons and trend monitoring through dashboards organized according to the rehabilitation results chain, spanning input, output, outcome, and efficiency domains. Training resources target both data entry staff and program managers, ensuring sustainable implementation across organizational levels.

The integration of the WHO RHIS – Rehabilitation module into Uganda’s National HMIS, as part of the DHIS2 platform, was executed through a structured, multi-step strategy, which is detailed in Section 3.5, and involved a comprehensive review process to align with national health priorities. After the review, Uganda’s MoH selected six core indicators from the original 15 for integration into the national DHIS2 platform. The finalized indicator set includes rehabilitation personnel density, rehabilitation service utilization, rehabilitation uptake, assistive products uptake, outreach programs uptake, and rehabilitation referral (**Figure 6**). While additional indicators, including waiting times, individualized care plans, and functional changes, will be captured in facility registers, their integration into digital reporting systems awaits future HMIS digitization. This phased approach reflects Uganda’s pragmatic balance between comprehensive data needs and implementation feasibility, establishing a foundation for progressive expansion of rehabilitation data capabilities within the national health information architecture.

**Figure 6:**
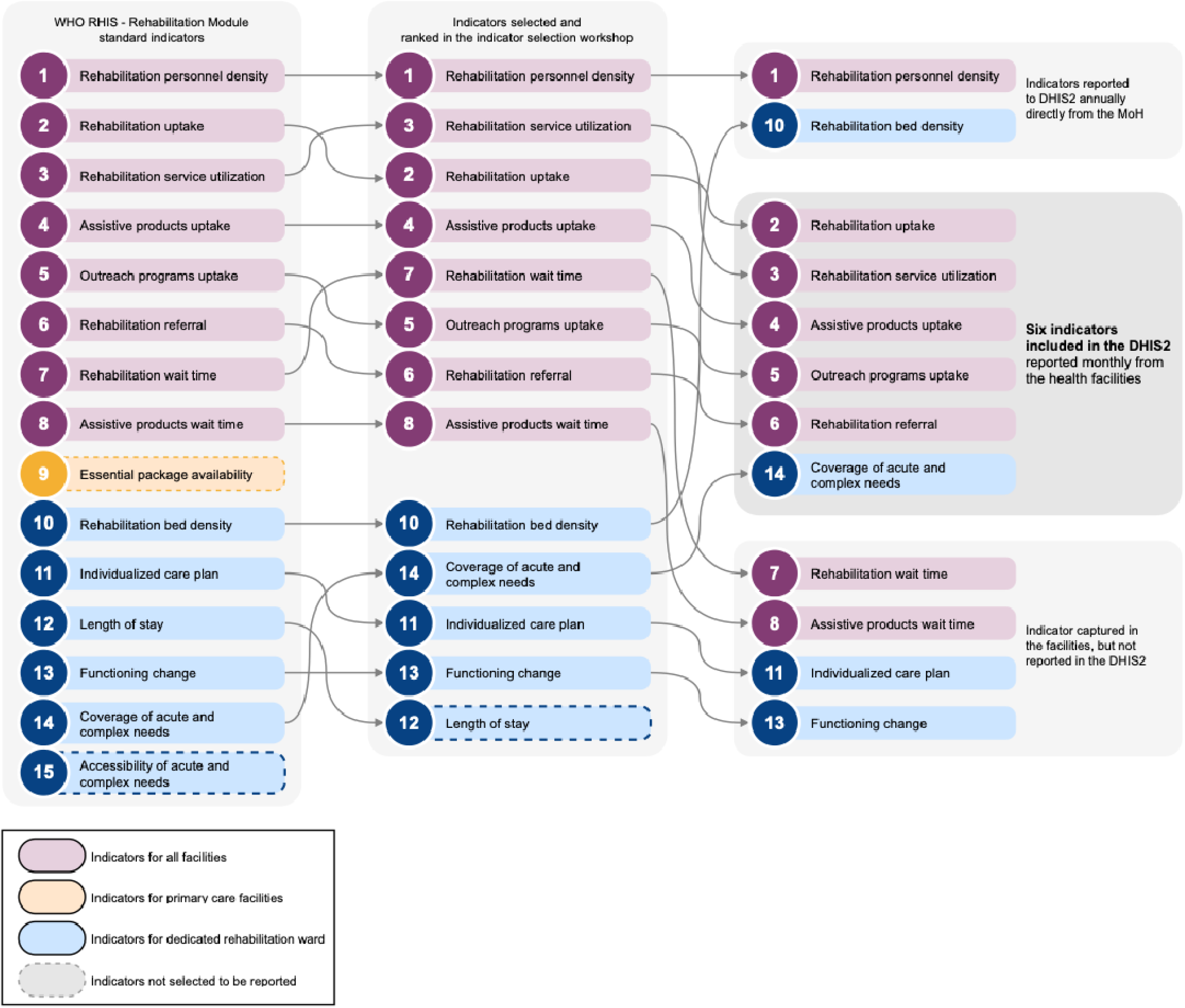
Indicator selection process and list of final rehabilitation indicators integrated into Uganda’s DHIS2.

### 3.5 Implementation Process of Integration

The integration of the WHO RHIS-Rehabilitation module into Uganda’s national HMIS followed a systematic, multi-phase approach spanning over two years from initial stakeholder engagement through active data collection (**Figure 7**). The process began with foundational stakeholder consultations involving the Ministry of Health, WHO, and ReLAB-HS in October 2022, strategically leveraging Uganda’s mid-term HMIS review as an opportunity to mainstream rehabilitation data within the national health information architecture. These consultative meetings established clear project objectives, roles, and responsibilities while securing critical buy-in from key government stakeholders and decision-makers. The timing proved advantageous, as the MoH was already exploring opportunities to enhance rehabilitation and assistive technology data collection within the DHIS2 platform during this review period. Early engagement emphasized the critical need for comprehensive rehabilitation data integration to support evidence-based decision-making, resource allocation, and strategic planning for rehabilitation services across Uganda’s health system.

**Figure 7:**
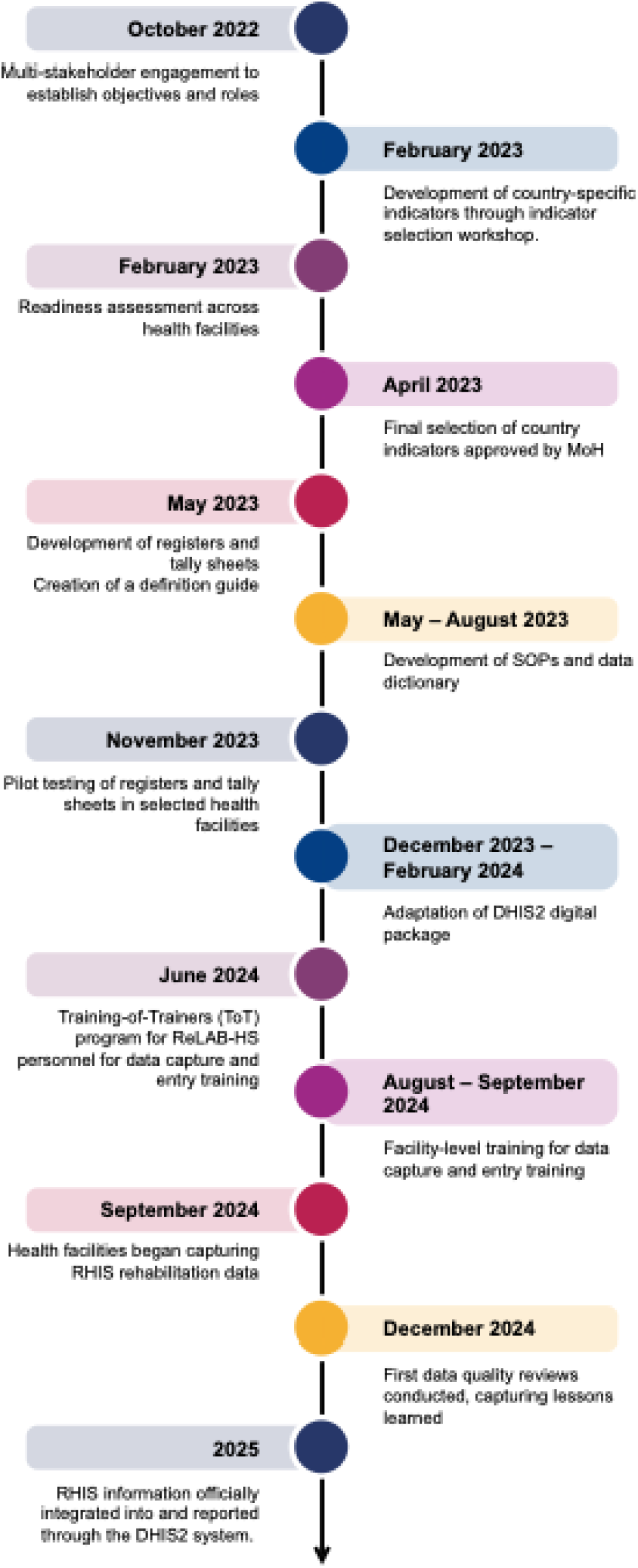
Timeline of integrating the WHO Routine Health Information System – Rehabilitation module into Uganda’s National Health Information System.

A comprehensive indicator selection workshop held in February 2023 at Jinja district convened 43 diverse stakeholders, including rehabilitation professionals (physiotherapists, occupational therapists, orthopedic technologists, audiologists), medical records specialists, HMIS officials, and representatives from the Disability and Rehabilitation Division. Over three days, participants systematically reviewed the WHO’s 15 standard RHIS-Rehabilitation indicators, evaluating each based on specific criteria, including usefulness for decision-making at all health system levels, facilitation of comprehensive analysis across results chain domains, ease of standardization, relevance for regional and global reporting, availability of variables in facilities, and limited data collection burden. The workshop employed structured breakout sessions and plenary discussions to achieve consensus, ultimately agreeing to include 13 of the 15 WHO-proposed indicators and ranking them according to national priorities. However, following submission to the MoH Health Information Systems division for final review, budgetary constraints and competing sectoral priorities led to the removal of seven indicators, leaving six core rehabilitation indicators for DHIS2 integration. This reduction highlighted the complex negotiation processes inherent in health system strengthening initiatives and the importance of early engagement with high-level decision-makers to minimize indicator attrition during implementation.

Parallel to indicator selection, a comprehensive readiness assessment was conducted across 12 strategically selected facilities, including two National Referral Hospitals and 10 Regional Referral Hospitals, starting in February 2022. The structured assessment employed WHO-developed checklists to evaluate human resources, rehabilitation service delivery capacity, data reporting systems, information use practices, and infrastructure availability. We have already presented some of the key findings in Sections 3.2 and 3.3. These findings informed subsequent capacity-building interventions and highlighted the need for substantial technical support during implementation.

The technical development phase from April through November 2023 involved intensive collaboration among MoH, WHO, and ReLAB-HS to modify existing HMIS tools and create new data collection instruments. The most significant innovation was the development of an independent outpatient rehabilitation register (HMIS REH 002), directed by MoH, which elevated the importance of rehabilitation data collection while enabling more detailed capture of rehabilitation-specific indicators. This register additionally incorporated the “Waiting Time” indicator, though this metric would be analyzed at the facility level rather than reported through DHIS2. Similarly, inpatient services were integrated through modifications to the existing inpatient register (HMIS IPD 003). Although indicators such as “Functioning Change” and “Individualized Care Plans” were captured, they were not immediately reported to DHIS2, pending the future digitization of inpatient systems at facilities with dedicated rehabilitation wards. The development process also produced comprehensive supporting materials, including updated data dictionaries, detailed definition guides, standard operating procedures, and individual reporting forms (HMIS 105 for outpatients, HMIS 108 for inpatients) used by HMIS focal persons for data entry. Pilot testing of revised tools across 18 health facilities during November 2023 enabled refinement based on user feedback and real-world implementation challenges.

Platform customization and capacity building commenced in December 2023 when the MoH Health Information Division, supported by the Health Information Systems Programme (HISP), began DHIS2 modifications to accommodate the finalized rehabilitation indicators. This technical adaptation occurred within an integrated approach, as part of the mid-term review of HMIS, encompassing multiple health sectors rather than as a standalone rehabilitation module. Next, in collaboration with WHO and ReLAB-HS central team, a comprehensive Training-of-Trainers program in June 2024 equipped MoH staff and ReLAB-HS technical advisors with skills to conduct facility-level orientations. The program co-developed training materials that addressed anticipated implementation challenges through role-playing exercises and scenario-based discussions. The ToT program proved particularly valuable as it not only developed training materials but also involved extensive discussions anticipating the types of questions trainers would encounter. The program involved role-playing and multiple scenarios considering local context and geographical variability. This thorough preparation enabled trainers to resolve many issues during actual implementation because they were well-prepared for various challenges. Following the ToT, these trained facilitators conducted regional training sessions among more than 100 HMIS focal points between August and September 2024, covering seven regions across Uganda’s 15 sub-regions.

Data collection officially commenced in September 2024 across 25 facilities, including four National Referral Hospitals, 17 Regional Referral Hospitals, and three district hospitals, covering 15 sub-regions of Uganda. The implementation faced initial challenges, including the simultaneous introduction of digital platforms at the point of care, which created confusion among staff accustomed to paper-based systems. A delayed DHIS2 launch generated resistance among health workers, and sustainability concerns arose regarding register printing and distribution, which depended on donor support. Nonetheless, high levels of enthusiasm among rehabilitation professionals and hospital management provided strong foundations for success, with one data personnel commenting that the new indicators solved longstanding challenges in identifying performance measures for rehabilitation professionals. Continuous technical support through WhatsApp groups enabled real-time problem-solving and knowledge sharing among trained personnel, while planned quarterly data quality reviews, beginning in December 2024, ensured adherence to protocols and progressive improvement in data accuracy. Although full online reporting through DHIS2 is scheduled for 2025, facilities began systematic data capture and storage to facilitate retrospective entry once the platform becomes fully operational.

## 4. DISCUSSION

This study examined the integration of the WHO RHIS-Rehabilitation module into Uganda’s national HMIS, documenting a comprehensive implementation process that successfully established standardized rehabilitation data collection across 25 health facilities. The integration achieved significant milestones, including the development of customized data collection tools, the capacity building of over 100 health workers, and the establishment of a systematic rehabilitation data capture system, which began in September 2024. However, the process also revealed substantial challenges across multiple implementation domains, including indicator reduction due to competing health sector priorities, infrastructure constraints, and workforce limitations. Using the CFIR, our analysis identified critical success factors and barriers that provide actionable insights for similar health system strengthening initiatives in LMICs (**Table 3**).

**Table 3:** Implementation challenges and opportunities across CFIR Domains for integrating the WHO Routine Health Information System – Rehabilitation module within Uganda’s DHIS2.

| Implementation Domains | Challenges | Opportunities |
| --- | --- | --- |
| **Individuals**<br>*refers to those involved in implementation, including MoH officials, rehabilitation professionals, data managers, and healthcare workers. It considers their knowledge, skills, attitudes, and the role of champions, training, and capacity-building in adopting the RHIS - Rehabilitation Module.* | Rehabilitation professionals faced confusion during the transition to the new indicator set due to a lack of communication from the MoH.<br>Staff shortages in facilities hindered consistent data collection and reporting.<br>Resistance to adopting new processes due to busy schedules and high patient workloads. | Motivated staff (e.g., rehabilitation professionals, HMIS focal points, and facility in-charges) viewed the new indicators as solutions for performance measurement. |
| **Inner Setting**<br>*focuses on Uganda's rehabilitation service delivery system, including referral hospitals and district facilities. It examines infrastructure, organizational culture, workflows, workforce capacity, internet connectivity, and the new module's compatibility with existing DHIS2 indicators and processes.* | The existing DHIS2 system lacked a dedicated set of rehabilitation indicators, limiting the capture of rehabilitation data.<br>Inconsistent reporting practices and data quality issues, such as missing data and outliers, were prevalent.<br>Lack of rehabilitation-specific services and providers at primary healthcare levels restricted data collection.<br>High patient loads disrupted training sessions and data collection processes in health facilities.<br>Limited infrastructure, such as space, computers, and internet access, hindered effective training and data collection. | On-site training enabled hands-on learning and allowed trainers to address real-world challenges.<br>Enthusiasm among rehabilitation professionals and data personnel ensured commitment to the new system.<br>Facility-level engagement supported advocacy for rehabilitation services and improved local ownership.<br>Digital transformation offered opportunities for streamlining data collection and improving data timeliness. |
| **Outer Setting**<br>*encompasses the broader context influencing Uganda's HMIS, including national policies, socioeconomic factors, donor influences, and international guidelines. It reflects how external pressures, the Ministry of Health, WHO, and partners shape rehabilitation data integration into DHIS2.* | Competing priorities, such as TB and HIV, limited the space available for new rehabilitation indicators.<br>Policy barriers delayed the integration of rehabilitation data into broader health information systems.<br>Limited government funding for printing and distributing registers hindered sustainability.<br>Fragmentation of rehabilitation and assistive technology data systems across different sectors (e.g., humanitarian settings, healthcare financing) | Leveraging the Ministry of Health's mid-term review of HMIS facilitated integration into national priorities.<br>Existing policies, like the Disability Act 2020, support building an enabling environment that ensures the inclusion of rehabilitation data into HMIS.<br>Cross-sector partnerships provided avenues to improve rehabilitation service delivery.<br>Increased advocacy for rehabilitation services elevated its status within national health policies. |
| **Innovation**<br>*refers to the characteristics of the RHIS - Rehabilitation module itself, including its features, structure, and purpose, which aims to standardize data collection on rehabilitation services and assistive technology within the DHIS2 system.* | Limited opportunity for integration of rehabilitation indicators in primary care settings since rehabilitation services are not available at primary care facilities. | The WHO RHIS - Rehabilitation module's standardized indicators enhanced data-driven decision-making and service monitoring.<br>Minimum adaptations to the RHIS - Rehabilitation module facilitated easier alignment with Uganda's health system.<br>Integrating rehabilitation data into digital platforms increased the visibility and importance of rehabilitation services.<br>The module's flexibility allowed future expansion to include more detailed indicators and service categories. |
| **Implementation Process**<br>*covers activities and strategies* | Some indicators were removed during the Ministry of Health's review due to | Collaboration with the Ministry of Health and WHO facilitated smooth navigation through the |
| *for integrating the RHIS - Rehabilitation Module into DHIS2, including stakeholder engagement, readiness assessments, indicator selection, tool development, and training. It emphasizes planning, execution, continuous support, feedback, and monitoring for sustainable implementation.* | budget constraints and priorities.<br>• Developing independent outpatient rehabilitation registers raised concerns about long-term funding and sustainability.<br>• Sustained funding was needed for printing registers, which relied on donor support.<br>• Decentralized training faced logistical issues due to limited space and high patient volumes in facilities.<br>• Initial feedback from the facilities revealed variations in reporting practices and inconsistencies in data capture. | integration process.<br>• Development of a detailed definition guide, standardized data interpretation, and enhanced reporting consistency.<br>• Role-playing and scenario-based training prepared trainers to address common challenges effectively.<br>• Training at health facilities provided direct engagement with hospital management, providing additional support.<br>• Continuous technical support, including quarterly data quality reviews, improved data accuracy and adherence.<br>• Establishing a WhatsApp group enabled real-time support and knowledge-sharing among trained staff. |

The implementation process demonstrated the critical importance of strategic timing and government ownership in health system reforms. Leveraging Uganda’s mid-term HMIS review created an opportune window for rehabilitation data integration, while strong MoH leadership proved instrumental in navigating bureaucratic challenges and maintaining implementation momentum. The reduction from 15 to 6 indicators during the MoH review process, while initially disappointing, highlighted the reality of resource constraints and competing health sector priorities that characterize many LMIC settings. This experience underscores the need for early engagement with high-level decision-makers and realistic expectation setting during the planning phase. The development of an independent outpatient rehabilitation register (HMIS REH 002), though innovative in elevating rehabilitation data visibility, also raised sustainability concerns regarding long-term funding for printing and distribution, which remained dependent on donor support.

Training and capacity building emerged as both a significant strength and an ongoing challenge throughout the implementation. The decision to conduct facility-based rather than centralized training proved highly effective in addressing real-world implementation challenges and fostering direct engagement with hospital management. This approach enabled trainers to provide contextualized support while advocating for additional institutional support from facility administrators. However, the decentralized training strategy also exposed infrastructure limitations, including inadequate training spaces, high patient volumes that disrupted sessions, and the complexity of managing simultaneous digital transformation initiatives at the point-of-care. The establishment of WhatsApp groups for ongoing technical support demonstrated the value of continuous engagement and real-time problem-solving, contributing to sustained data quality improvements and staff confidence in using the new systems.

The integration revealed fundamental structural challenges within Uganda’s rehabilitation service delivery system that extend beyond data collection to core service availability and workforce capacity. The concentration of rehabilitation services primarily at referral hospital levels, combined with critical workforce gaps, particularly in speech therapy, audiology, and specialized medical rehabilitation, limited the potential scope of data collection to higher-tier facilities. This structural reality meant that the WHO RHIS module could not be implemented at primary healthcare levels where rehabilitation-specific providers are absent, representing a significant limitation for comprehensive health system coverage. Nevertheless, the enthusiasm demonstrated by rehabilitation professionals and hospital management, exemplified by one data staffer’s comment that “we have been struggling to identify performance indicators for rehabilitation professionals; these indicators have solved our problem,” suggests strong organizational readiness for systematic performance measurement and quality improvement initiatives.

This study employed a comprehensive case study methodology that triangulated multiple data sources, including organizational reports, rapid readiness assessments, national household surveys, and policy documents, to provide detailed implementation insights. While this approach may lack the quantitative rigor, it offers robust triangulation and captures the complex, contextual factors that influence health system interventions in real-world settings. A significant limitation was our inability to report comprehensive data quality assessment results due to premature funding withdrawal from the donor organization, which prevented systematic evaluation of data accuracy and completeness across participating facilities. However, this constraint paradoxically provided valuable insights into the sustainability challenges facing health system strengthening initiatives in donor-dependent environments, highlighting the critical importance of securing long-term financial commitments for successful implementation and evaluation of complex health information system reforms. The application of an implementation science framework, specifically the CFIR, enabled systematic identification and categorization of barriers and facilitators across multiple domains, providing structured insights that enhanced both our understanding of complex implementation dynamics and the generalizability of findings. Lastly, our case design was appropriate as Uganda’s experience represented a unique, information-rich case of systematic integration of rehabilitation data into a national HMIS, providing insights that could inform similar initiatives in other LMICs seeking to strengthen health information systems for previously neglected health services.

## 5. CONCLUSION

The integration of the WHO RHIS – Rehabilitation module into Uganda’s National HMIS signifies a critical milestone in strengthening rehabilitation data collection and management. By incorporating standardized indicators, enhancing digital reporting through DHIS2, and supporting capacity-building efforts, this activity lays a strong foundation for evidence-based decision-making for the rehabilitation sector. Despite challenges such as infrastructure limitations, workforce shortages, and competing health priorities, the collaborative approach between the MoH, WHO, and ReLAB-HS has demonstrated the importance of stakeholder engagement, contextualized training, and ongoing technical support. Lessons from this implementation underscore the importance of sustained investment, continuous refinement of data systems, and enhanced policy integration to ensure long-term success. Moving forward, expanding rehabilitation data collection to lower-tier facilities, enhancing digitalization, and addressing resource gaps will be critical. By strengthening rehabilitation information systems, Uganda is advancing its commitment to universal health coverage and ensuring equitable access to essential rehabilitation services for all.

## Data Availability

All data produced in the present study are available upon reasonable request to the authors

## Glossary

Assistive Technology (AT): Devices or products—such as wheelchairs, hearing aids, or prosthetics—that enhance an individual’s ability to function and participate in daily activities.
Consolidated Framework for Implementation Research (CFIR): A multi-domain implementation science framework that structures analysis of contexts, processes, and determinants influencing health system interventions.
Data Dictionary: A reference document defining each data field, indicator, and reporting variable used within HMIS and associated registers.
DHIS2 (District Health Information System 2): An open-source digital platform used for health data management and reporting at national and subnational levels; serves as Uganda’s backbone for electronic HMIS.
Health Management Information System (HMIS): An integrated system for routine collection, processing, and reporting of health service data at facility and national levels, used for planning, monitoring, and management.
Implementation Science: The interdisciplinary study of methods to promote the systematic uptake of research findings and evidence-based practices into regular use by health systems.
Rapid Readiness Assessment: A structured evaluation conducted prior to implementation to assess health facility infrastructure, workforce capacity, and digital preparedness for new systems.
Rehabilitation: A set of interventions designed to optimize functioning and reduce disability in individuals with health conditions, including both medical and community-based services.
Standard Operating Procedures (SOPs): Documented, step-by-step instructions to ensure standardized processes in data collection, entry, and reporting within facility and national systems.
Universal Health Coverage (UHC): A health system goal to provide all individuals access to essential health services—including rehabilitation—without financial hardship.
WHO RHIS-Rehabilitation Module: A standardized data collection and reporting toolkit developed by the World Health Organization for integrating rehabilitation indicators into national health information systems, including DHIS2.

## Funding Sources

This publication is made possible by the support of the American people through the United States Agency for International Development (USAID) through the Learning Acting Building for Rehabilitation in Health Systems (ReLAB-HS) project (Grant No. 135966). The contents are the sole responsibility of ReLAB-HS and do not necessarily reflect the views of USAID or the United States Government.

## Ethical Approval

This study (IRB No. 00014471) was received ethical approval by the Johns Hopkins School of Public Health (JHSPH) Institutional Review Board

## Notes

### Competing Interest Statement

The authors have declared no competing interest.

### Funding Statement

This study was funded by USAID

### Summary of Updates

Removing personal details from the title page

